# Non-random Rotation Matching Algorithm

**DOI:** 10.1101/2023.08.28.23294718

**Authors:** Roshan Lodha, Neil Mehta, Craig Nielsen

**Affiliations:** Cleveland Clinic Lerner College of Medicine, Cleveland, OH

**Keywords:** Medical Education, Linear Sum Assignment

## Abstract

Assigning clerkship positions to third-year medical students is crucial for their professional development. However, the process is challenging due to limited positions and the requirement to align students with suitable clerkships matching their interests and career goals. In this paper, we explain our approach of treating clerkship assignments as a linear-sum optimization problem. This optimizes position assignments, minimizing costs. We believe this approach could enhance the clerkship assignment process and improve medical students’ learning.

## 1 Introduction

Third-year medical students engage in core rotations covering internal medicine, surgery, pediatrics, obstetrics, gynecology, neurology, psychiatry, and family medicine. Elective rotations often necessitate prior completion of core rotations; for instance, to take the orthopedics elective, students must first finish the core surgery rotation. This makes students inclined to have preferences regarding the sequence of their rotation assignments.

Currently, rotation matching relies on a stratified-random approach, consuming significant faculty time and introducing bias through stratification criteria. More critically, this method does not permit students to express their desired clerkship order. Here, we propose a student-centered rotation assignment algorithm aimed at achieving the best pairing of students and their preferred order. Importantly, our approach allows students to individually determine the cost of an unfavorable assignment, granting them greater control compared to a ranked preference list.

## 2 Methods

### 2.1 Problem Formulation and Encoding

We recast the optimal rotation order challenge as a minimum-cost assignment issue. Each student can select from *k* rotation orders, each with an associated cost (see Section 2.1.1). These costs were then utilized to construct an *n* × *k* matrix.

#### 2.1.1 Determining Costs

Students received *b* “beans” and allocated them to the rotation orders at their discretion. Null submissions were allocated *b/k* beans to each rotation. All responses were scaled so that the sum of beans assigned by each student equated to *b*. The assigned beans for each rotation order were subsequently converted into a cost by subtracting the bean count from *b*. Hyperparameter optimization was applied to ascertain the optimal bean count for a specific application.

#### 2.1.2 Linear Cost Alternative

For enhanced applicability, our algorithm accommodates a ranked-preference-based assignment optimizer along with a beans-to-rank conversion tool. However, in practice, this tool was not utilized (see Section 5).

### 2.2 Algorithm Design

#### 2.2.1 Matrix Padding

Linear sum optimization necessitates a wide or square matrix. Therefore, we introduced phantom students without rotation order preferences until the number of rows became a multiple of *k*. Subsequently, we expanded the matrix to a width of ⌈*n*/*k*⌉, resulting in a square matrix. The row order was randomized to eliminate submission time bias in determining rotation order preference.

#### 2.2.2 Linear Sum Optimization

We calculated the optimal rotation order by conducting linear sum optimization on the padded, square cost matrix using Python (SciPy 1.9.3, Python 3.9.6).

### 2.3 Error Testing

To gauge the rotation assignment’s performance, we introduced a novel error metric, *δ*, defined as

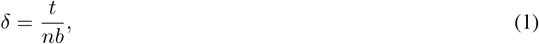

where *t* represents the total cost. *δ* is a real number in the range [0, 1], with a lower value indicating closer alignment with the optimal outcome.

## 3 Worked Example

Consider the following bean assignments matrix *M* for 7 students and 4 clerkship orders.

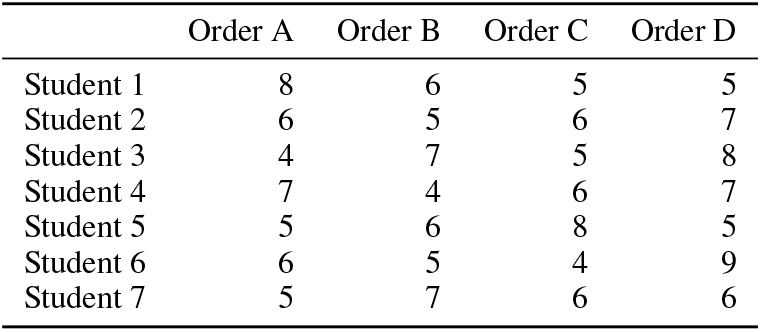

|  | Order A | Order B | Order C | Order D |
| --- | --- | --- | --- | --- |
| Student 1 | 8 | 6 | 5 | 5 |
| Student 2 | 6 | 5 | 6 | 7 |
| Student 3 | 4 | 7 | 5 | 8 |
| Student 4 | 7 | 4 | 6 | 7 |
| Student 5 | 5 | 6 | 8 | 5 |
| Student 6 | 6 | 5 | 4 | 9 |
| Student 7 | 5 | 7 | 6 | 6 |

The sum of each row (the total number of beans assigned by each student) is 24. Thus, the cost associated with a given order can be given by 24 − *M*_*i,j*_. The number of phantom students is given by *k* − (*n* mod *k*), or 1 in this case. Following cost conversion and phantom student insertion, the resulting matrix is given by *M* ′.

This cost matrix is then tiled to form a square matrix, *M*′′.

The Jonker & Volgenant algorithm is run on *M*′′ resulting in the final student assignments:

- Student 1: Order C
- Student 2: Order D
- Student 3: Order A
- Student 4: Order B
- Student 5: Order A
- Student 6: Order C
- Student 7: Order B

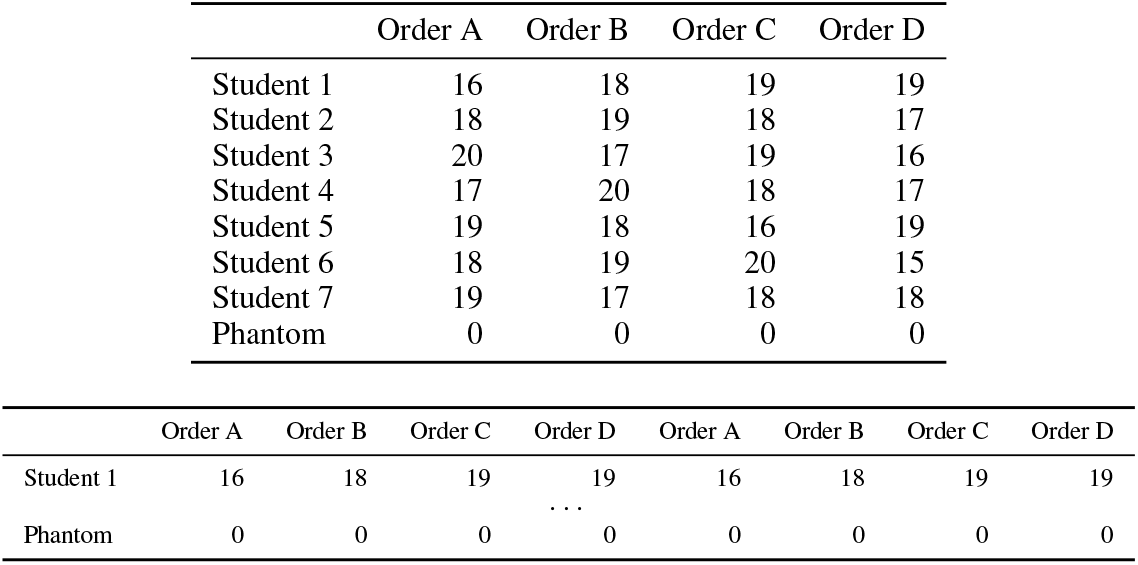

## 4 Results

### 4.1 Simulations

In simulations, we found that the optimal number of beans was highly variable. In general, a minimum number of beans minimized error (Figure 1a) given a uniform-random cost matrix.

**Figure 1.**
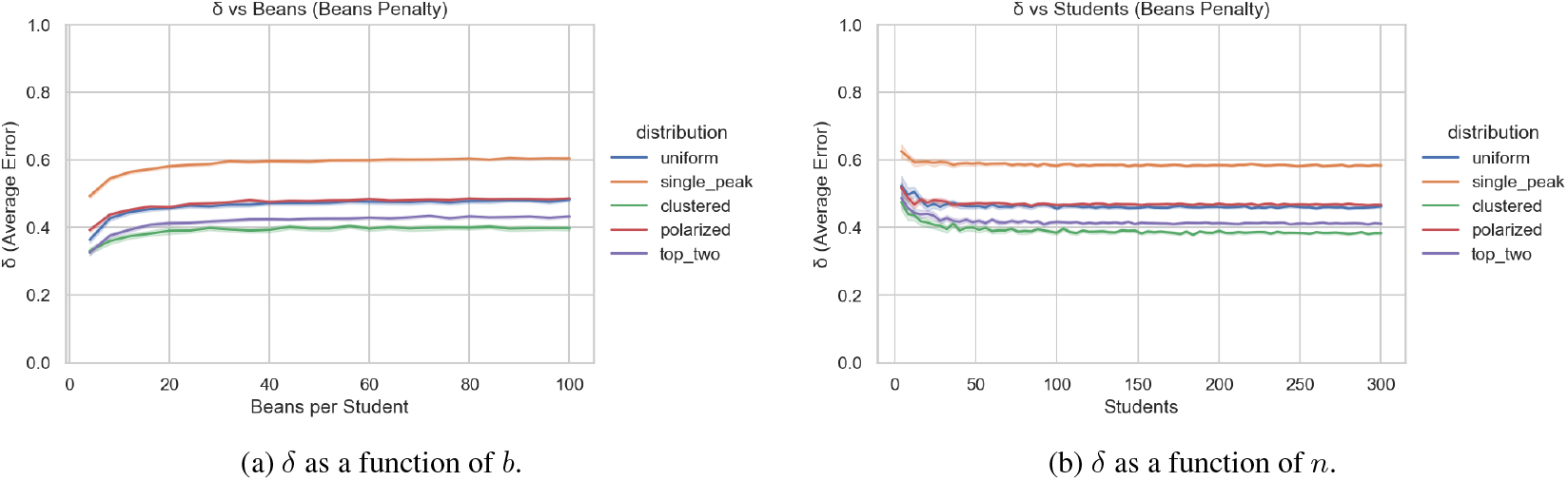
*δ* tends to increase as the number of beans increases but tends to decrease as the number of students increases.

As the number of students increases, the total *δ* error decreased exponentially (Figure 1b).

### 4.2 Practical deployment

#### 4.2.1 Definition of Parameters

We asked *n* = 77 students to assign *b* = 24 beans to any permutation of *k* = 4 possible rotation orders (Table 1). We employed a maximum number of *b* = 24 beans per student to allow for integer divisions of beans into preferences. We chose 24 beans as it is *k*! allowing for integer divisions into each possible category.

**Table 1:** Rotation order definitions.

| Option | Clerkship 1 | Clerkship 2 | Clerkship 3 | Clerkship 4 |
| --- | --- | --- | --- | --- |
| 1 | LAB | TBC2 | TBC3 | TBC1 |
| 2 | TBC2 | LAB | TBC1 | TBC3 |
| 3 | TBC3 | TBC1 | LAB | TBC2 |
| 4 | TBC1 | TBC3 | TBC2 | LAB |

#### 4.2.2 Initial Deployment

Real-world behavior in rotation order selection is poorly modeled by a uniform distribution. Specifically, we found that rotation order 4 *>* rotation order 3 *>* rotation order 2 *>* rotation order 1 (Table 2). This led to several students receiving a deeply unfavorable (last choice) clerkship order option. Further testing must be done to determine how the beans-count affects the error under various sampling distributions. We hypothesize that in real-world deployment, increasing the number of beans would decrease the error due to sampling skew and a maximal difference between costs for a given student.

**Table 2:** Summary statistics of beans assignment by clerkship order option.

| Option | Order | Beans Assigned | Number All In |
| --- | --- | --- | --- |
| 1 | LAB–TBC2–TBC3–TBC1 | 271 | 2 |
| 2 | TBC2–LAB–TBC1–TBC3 | 475 | 13 |
| 3 | TBC3–TBC1–LAB–TBC2 | 859 | 26 |
| 4 | TBC1–TBC3–TBC2–LAB | 490 | 15 |

**Table 3:** Performance metrics of the algorithm on historical student cohorts.

| Year | Students (N) | First Choice % | Top 2 % | Avg. Penalty (Beans) |
| --- | --- | --- | --- | --- |
| 2023 | 77 | 72.7% | 87.0% | 6.26 |
| 2024 | 77 | 80.5% | 92.2% | 4.61 |
| 2025 | 75 | 78.7% | 92.0% | 5.32 |

#### 4.2.3 Subsequent Deployment

To validate the efficacy of the Non-random Rotation Matching Algorithm (NRMA) in a real-world setting, we analyzed historical preference data from three consecutive academic years (2023–2025). The dataset comprised responses from 229 students (77 in 2023, 77 in 2024, and 75 in 2025).

The algorithm demonstrated consistent performance across all cohorts, achieving a mean first-choice assignment rate of 77.3% and an average penalty (cost) of 5.40 beans per student (0 indicates a first-choice assignment).

Figures 2a–c summarize how students distributed their 24 beans across the four rotation-order options in each cohort year.

**Figure 2.**
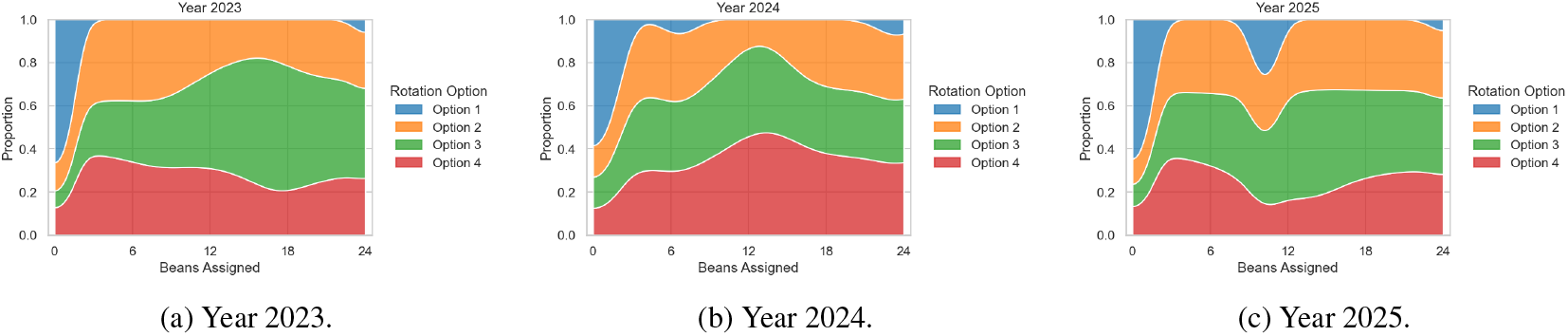
Distribution of beans assigned to each rotation-order option by year.

Figures 3a–c show the distribution of bean penalties (cost) per student for each cohort year.

**Figure 3.**
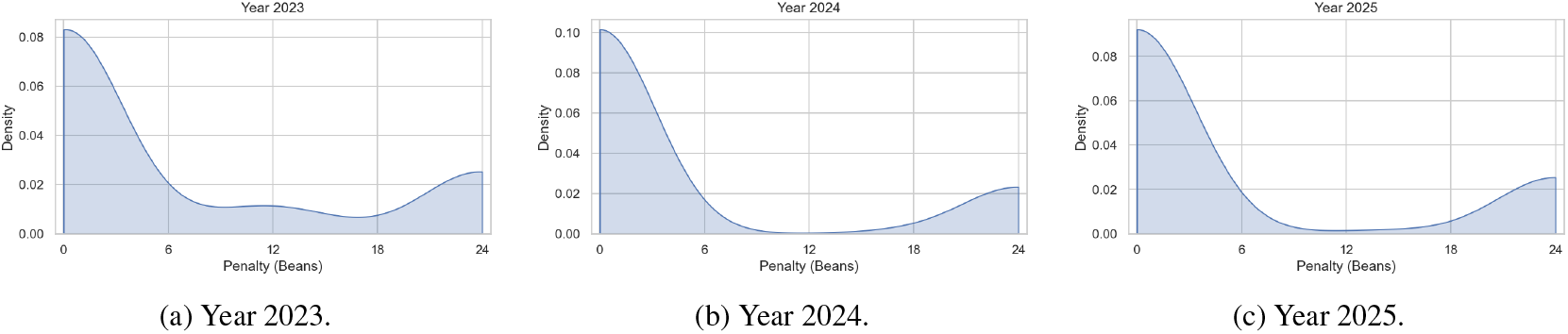
Distribution of bean penalty per student by year.

## 5 Discussion

### 5.1 Key Findings

#### 5.1.1 Optimal and Complete

In our problem, optimality was defined as a rotation order assignment in which no single swap would benefit all students involved in the swap. Completeness was defined as both an equal number of students assigned to each rotation order as well as all students being assigned to exactly 1 rotation order. In the case that the number of students was not 0 in the moduli space *k*, completeness was defined as a difference of no more than 1 student between the most filled and least filled rotation groups. Linear sum optimization provides an optimal solution by definition. Completeness was ensured by matrix padding.

#### 5.1.2 Optimal Student Strategy

In the context of student rotations, the influence of student-determined cost penalties and the variable popularity of different rotations introduces an opportunity for strategic application of game theory principles. Consider the data presented in Table 2. In this scenario, a plurality of students preferred rotation option 3, while only a single student preferred option 1. By definition, a maximum of 1/4 of students could be accommodated with their first-choice rotation. Thus, the vast majority of students allocated all their beans to option 3, thereby maximizing their chances of obtaining it. However, since more than 1/4 of the students adopted this strategy several students were randomly assigned to alternative rotations. Given the relative unpopularity of option 1, it is likely that a majority of these students would be assigned to it. Consequently, students may find it advantageous to consider assigning their resources to their second-choice rotation, accepting the possibility of conceding their first-choice.

Our algorithm allows for easy modification and elimination of this aspect by having students rank their preferences followed by deterministic assigning a cost penalty to an unfavorable rotation order without the students’ consultation.

Simulations of this method similarly showed decreased error with increased number of students, indicating that error is roughly constant under a linear penalty regime. In practice, this was not used to allow finer control over students’ choices.

### 5.2 Future Directions

#### 5.2.1 Skewed costs

Applying a weight to the cost matrix can skew the results to avoid assigning students to their last-choice preference. For example, adding an exponential penalty would more significantly penalize rotation orders with fewer beans, skewing the optimal result away from those sets of solutions. Our algorithm allows for easy modification of a cost matrix. Hyperparameter optimization should be used to determine the best cost penalty function for a given application.

#### 5.2.2 Adding distance penalties and car-share benefits

Within each rotation, students can be placed at several sites. Suburban hospital campuses pose an additional cost to students in the form of travel. Future iterations of a non-random rotation matching algorithm can modify the cost function based on the distance a student has to travel to a given rotation. An example implementation could be to recursively run the algorithm within each rotation assignment using the distance traveled in miles as the of a rotation. Similarly, students often live with another medical student. To encourage carpooling, the cost function can be further modified to increase the odds that two students are placed in the same rotation.

#### 5.2.3 Unequal rotation sizes

While our usage mandated an equal number of students in each rotation, updating the algorithm for unequal distributions is as simple as modifying the tiling function to include more repetitions of options that can accommodate a higher number of students.

## 6 Conclusion

Assigning clerkship positions to third-year medical students is a crucial step in their training. However, the process is complex due to limited positions and the need to match students with suitable clerkships. This study proposed a new way to approach this problem, treating it as a linear-sum optimization challenge. By using this method, we aimed to distribute clerkship positions to students in a way that minimizes costs.

The potential impact of this approach is significant. It could improve the efficiency of clerkship assignments and enhance the learning experiences of medical students. As institutions strive to make clerkship placements more effective, this new approach offers a promising avenue. It could lead to better outcomes, ensuring a more productive and valuable educational journey for future medical professionals.

## Data Availability

All data produced are available online at https://github.com/roshanlodha/NRMA.

## Acknowledgments

This was supported part by the Cleveland Clinic Lerner College of Medicine of Case Western Reserve University. Deployment of this project to the first medical student batch was helped by Dr. Neil Mehta and Dr. Craig Nielsen of the Cleveland Clinic.

## Notes

### Competing Interest Statement

The authors have declared no competing interest.

### Clinical Protocols

https://github.com/roshanlodha/NRMA

### Funding Statement

This study did not receive any funding.

### Summary of Updates

included historical data analysis; improved stress testing to include various distributions; updated figures

